# Prevalence and Risk Factors of Hypertension Among Young Adults (26-35 Years Old) in Indonesia: Analysis of Secondary Data from Riskesdas 2018

**DOI:** 10.1101/2024.06.10.24308678

**Authors:** Laluna Rachma Putri, Arulita Ika Fibriana, Mahalul Azam

## Abstract

**Background:** Worldwide, there are 54% adults diagnosed with hypertension. The prevalence of hypertension increased, based on the basic health survey (Riset Kesehatan Dasar; RISKESDAS) in 2013, the prevalence of hypertension was 25.8%, while the on the RISKESDAS 2018 data increased to 34.1%. This research aimed to find out the risk factors of hypertension among young adults in Indonesia using RISKESDAS 2018 data.

**Method:** The research design approach observational quantitative by cross-sectional design. This research used secondary data from RISKESDAS 2018 with a total of 140,073 subjects aged 26-35 years. All subjects who fulfil the inclusion criteria will be analyzed (n=78,000). Data analysed by chi-square test and binary logistic regression.

**Results:** The result of research exposed gender (p<0.001), marital status (p<0.001), education levels (p<0.001), working status (p<0.001), BMI (p<0.001), diabetes mellitus (p<0.001), emotional mental disorder (p<0.001), and instant food consumption (p<0.001) has a significant consequence of hypertension.

**Conclusions:** The results of research can be used as a consideration for creating programs and policies to control risk factors or hypertension among young adults in Indonesia. Promotive and preventive policies programs related to hypertension must reach the community to reduce hypertension morbidity and mortality rates.

## Introduction

Non-communicable diseases (NCDs) are not caused by viruses, germs, or bacteria but by daily lifestyle. One of the NCDs that threatens human health is hypertension (Minasari et al., 2022). Hypertension is a circulatory system disorder that increases blood pressure (Azinar et al., 2019). The number of hypertension patients continues to increase every year. In 2025, there will be 1.5 billion people with hypertension. Every year, it is estimated that there will be 9.4 million deaths due to hypertension and its complications (World Health Organization, 2023). Worldwide, there are 54% of adults diagnosed with hypertension (Global Health Observatory, 2023). An estimated 46% of adults with hypertension are unaware of their condition (World Health Organization, 2023). The prevalence of hypertension increased, based on the basic health survey (Riset Kesehatan Dasar; RISKESDAS). In 2013, the prevalence of hypertension was 25.8%, while the RISKESDAS 2018 data increased to 34.1%. There are a plenty of unreported cases; merely 1/3 of hypertension cases in Indonesia are diagnosed (Rokom, 2021).

Hypertension affected economic losses, productivity decline, and the emergence of complications. Noteworthy, hypertension can cause complications in diabetes mellitus, making it a considerable risk factor for 39 cardiovascular diseases (Azam et al., 2023). Hypertension patients have a 2.6 times greater risk of developing coronary heart disease compared to non-hypertensive individuals (Amisi et al., 2018). Hypertension patients lose productivity without controlling for health risks and chronic conditions (MacLeod et al., 2022). Hypertension medication ranks first in terms of the highest cost in Indonesia, particularly IDR 12.1 trillion (Social Security Agency of Health, 2017).

The majority of young adults with hypertension are ignorant of abnormally high blood pressure and acknowledge the impact when they get older. There is a long-term impact of early-life risk factors of hypertension in young adults (Gooding et al., 2014). Hypertension patients aged below 40 years have low awareness, low diagnosis, and poor blood pressure control (Johnson et al., 2014). The prevalence of undiagnosed hypertension in men aged 25 to 34 years is 55% and 44% in women aged 25 to 34 years. The prevalence of undiagnosed hypertension in young adults is higher than in people aged 75 years or over (17% in men and 21% in women) (Lacobucci, 2023).

We must undertake preventive efforts against hypertension among young adults. It is essential to identify and maintain modifiable risk factors for hypertension. This study aimed to provide information on the risk factors for hypertension among young adults based on the results of basic health research in 2018 in Indonesia. The results of the analysis are expected to impact suitable hypertension prevention in Indonesia. Efforts against hypertension prevention positively impact the health of each individual and community welfare, reducing costs for hypertension medicine and reducing the disadvantage of disease caused by hypertension.

## Methods

### Design and Study Population

This research is an observational quantitative approach cross-sectional design. This study utilized secondary data from the basic health survey (Riset Kesehatan Dasar; RISKESDAS) 2018. RISKESDAS is a population health study conducted by the Agency for Health Policies Development, Ministry of Health of the Republic of Indonesia. RISKESDAS aims to monitor indicators of public health and indicators of public health services. RISKESDAS provides information on key health indicators such as health status, nutritional status, environmental health, health behaviors, and various aspects of health care. RISKESDAS data provides health data at district/city and national levels. This study population included subjects aged 26–35 who were interviewed by the RISKESDAS 2018 enumerator, with a total of 140,073 subjects aged 26–35. Of the 140,073 subjects, 78,300 met the criteria to be analyzed (Figure 1).

**Figure 1.**
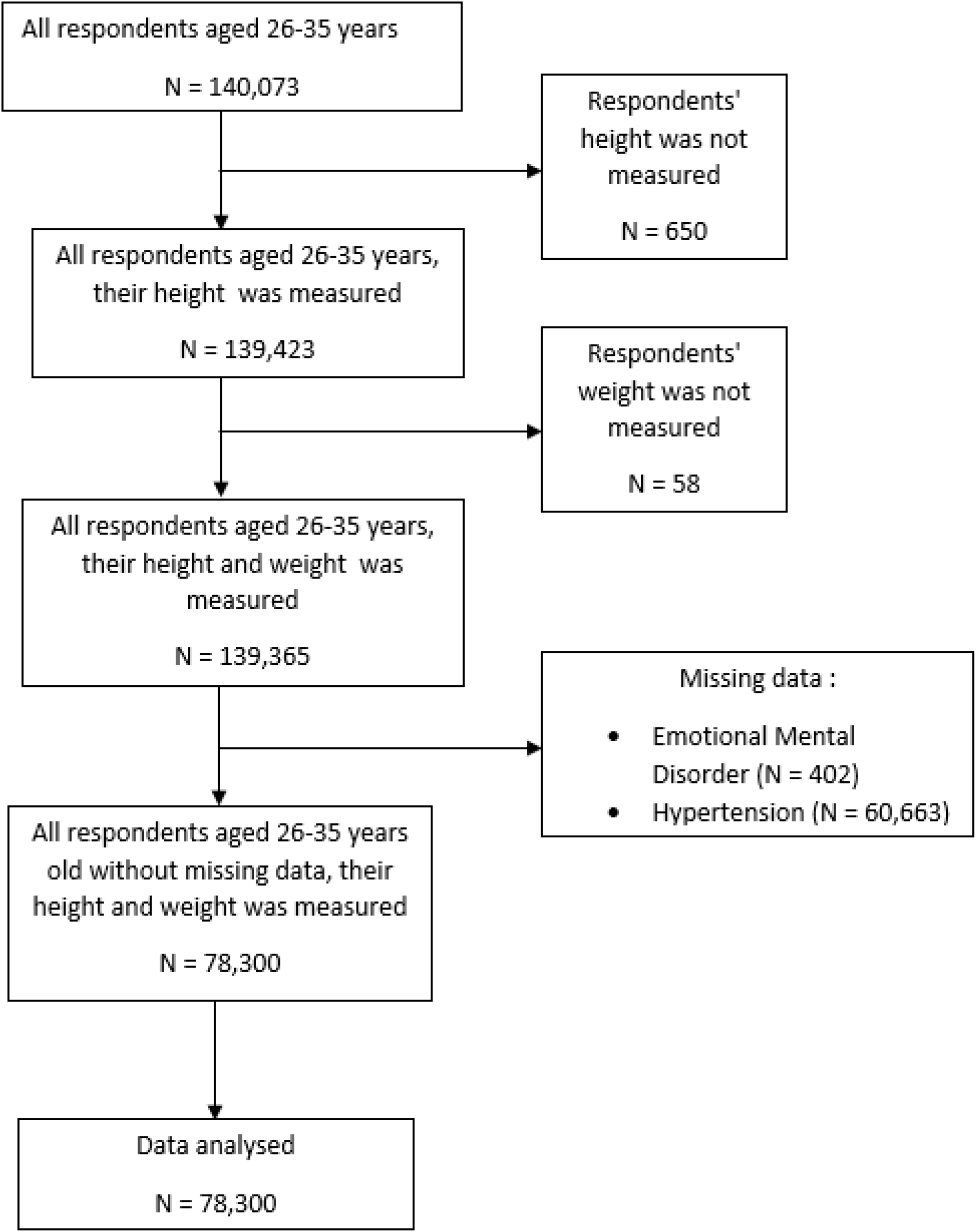
Sampling flow

### Data Collection

This study met the ethical criteria by Universitas Negeri Semarang of No.169/KEPK/FK/KLE/2024. The Agency for Health Policies Development, Ministry of Health of the Republic of Indonesia, has approved the use of RISKESDAS data for study purposes. This study has received permission to utilize, exploit, and process data in scientific studies. The subjects used in this research were subjects who already measured their weight and height; there were also no missing data.

This study used certain parameters from RISKESDAS 2018, which were analyzed in this research. These include sociodemographic data, hypertension status, diabetes mellitus status, emotional mental disorder status, smoking status, physical activity status, alcohol consumption, body mass index, salt food consumption, fat food consumption, and instant food consumption.

### Statistical Analysis

The characteristics of the subjects were categorical data, so data was presented in a proportion table. This study performed a chi-square test to find out the risk factors associated with hypertension among young adults in Indonesia. The p-values <0.05 reflected as statistically significant. This study involved multivariate analysis using binary logistic regression for all parameters with p-value <0.25 in the chi-square test. The statistical analysis was accomplished in this study using IBM SPSS Statistics software (for Windows, Version 22.0, Armonk, NY: IBM Corp.).

## Result

### Subjects’ Characteristics

Of the 140,073 subjects aged 26-35, 73,800 fulfilled the criteria and were included in further analysis. Among these, 3,225 (4.1%) young adults in Indonesia suffered from hypertension, whereas 75,075 (95.9%) did not. Table 1 explains the characteristics of these subjects.

**Table 1.** The characteristics of study subjects.

| **Variables** | **Frequencies**<br>**(N)** | **Percentages**<br>**(%)** |
| --- | --- | --- |
| **Gender** |  |  |
| Male | 27227 | 34.8 |
| Female | 51073 | 65.2 |
| **Marital Status** |  |  |
| Unmarried | 8398 | 10.7 |
| Married | 67895 | 86.7 |
| Widowed | 2007 | 2.6 |
| **Residence Area** |  |  |
| Urban | 34367 | 43.9 |
| Rural | 43933 | 55.1 |
| **Education Level** |  |  |
| Low | 37482 | 47.9 |
| High | 40818 | 52.1 |
| **Working Status** |  |  |
| No | 26342 | 33.6 |
| Yes | 51958 | 66.4 |
| **Body Mass Index** |  |  |
| BMI≥25 | 31430 | 40.1 |
| BMI<25 | 46870 | 59.9 |
| **Diabetes Mellitus** |  |  |
| Yes | 274 | 0.3 |
| No | 78026 | 99.7 |
| **Emotional Mental Disorder** |  |  |
| Yes | 7705 | 9.8 |
| No | 70595 | 90.2 |
| **Smoking** |  |  |
| Yes | 22,45 | 28.4 |
| No | 56055 | 71.6 |
| **Physical Activity** |  |  |
| No | 7067 | 9.0 |
| Yes | 71233 | 91.0 |
| **Alcohol Consumption** |  |  |
| Yes | 3540 | 4.5 |
| No | 74760 | 95.5 |
| **Salt Food Consumption** |  |  |
| Frequent (≥3-6 times/week) | 32153 | 41.1 |
| Rarely (<3-6 times/week) | 48147 | 58.9 |
| **Fat Food Consumption** |  |  |
| Frequent (≥3-6 times/week) | 47346 | 60.5 |
| Rarely (<3-6 times/week) | 30954 | 39.5 |
| **Instant Food Consumption** |  |  |
| Frequent (≥3-6 times/week) | 21864 | 27.9 |
| Rarely (<3-6 times/week) | 56436 | 72.1 |
| **Hypertension** |  |  |
| Yes | 3225 | 4.1 |
| No | 75075 | 95.9 |

**Table 2.**
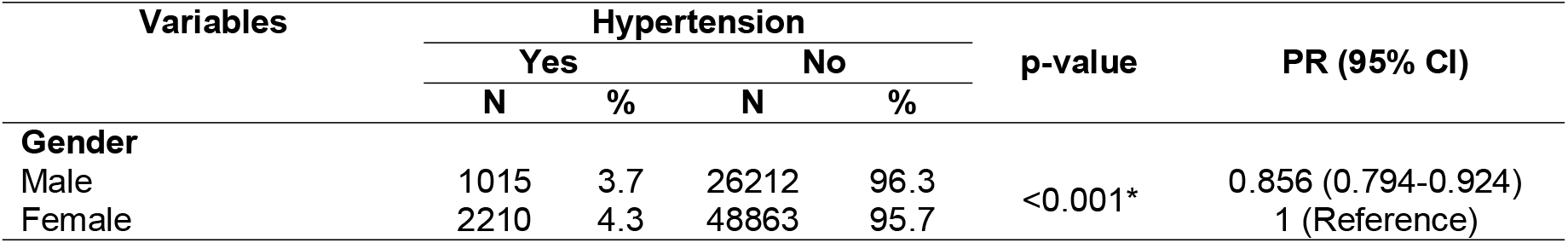
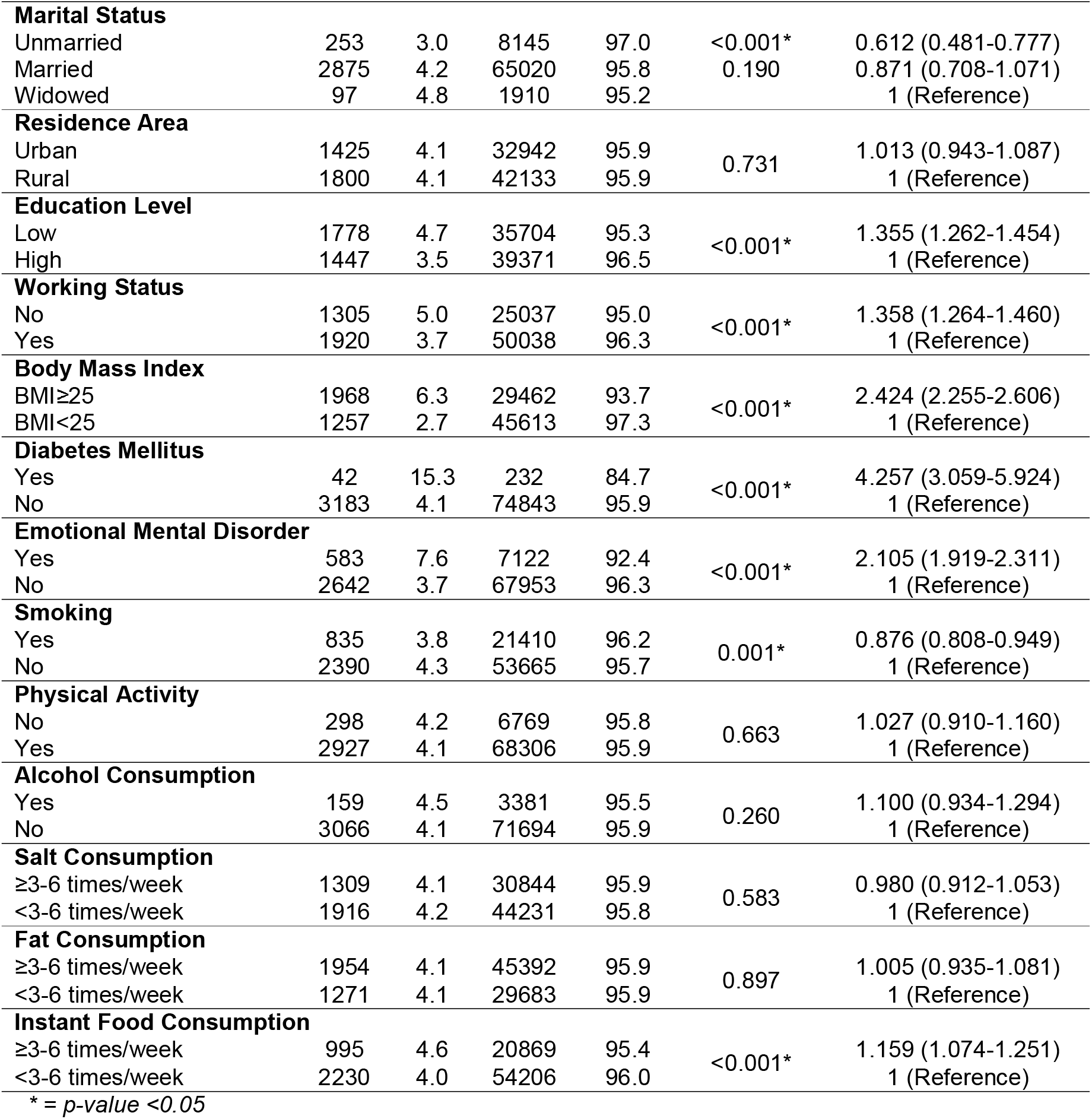
The characteristics of study subjects based on hypertension status.

### Risk Factors Associated to Hypertension among Young Adults (26-35 years) in Indonesia

Based on the result of the chi-square test (Tabel 2), some parameters show p-value <0.005; it refers to evidence of influence hypertension among young adults, i.e. gender (PR=0.856; 95% CI: 0.794-0.924), education levels (PR=1.355; 95% CI: 1.262-1.454), marital status (PR=0.612; 95% CI: 0.481-0.777), working status (PR=1.358; 95% CI: 1.264-1.460), body mass index (PR=2.424; 95% CI: 2.255-2.606), diabetes mellitus (PR=4.257; 95% CI: 3.059-5.924), emotional mental disorder (PR=2.105; 95% CI: 1.919-2.311), smoking (PR=0.876; 95% CI: 0.808-0.949), and instant food consumption (PR=1.159; 95% CI: 1.074-1.251). Variables with a p-value <0.25 will undergo continued analysis by binary logistic regression. Table 3 shows the final regression model results.

**Table 3.** Binary logistic regression of hypertension risk factors among young adults subject.

| Variables | B | S.E | Wald | p-value | PR (95% CI) |
| --- | --- | --- | --- | --- | --- |
| Gender | 0.203 | 0.046 | 19.368 | <0.001* | 1.225 (1.119-1.341) |
| Marital Status | -0.341 | 0.125 | 7.434 | 0.006* | 0.711 (0.556-0.908) |
| Education Levels | 0.251 | 0.037 | 45.177 | <0.001* | 1.286 (1.195-1.384) |
| Working Status | 0.237 | 0.043 | 30.930 | <0.001* | 1.267 (1.166-1.377) |
| BMI | 0.905 | 0.038 | 573.050 | <0.001* | 2.472 (2.295-2.662) |
| Diabetes Mellitus | 1.305 | 0.173 | 57.182 | <0.001* | 3.688 (2.630-5.173) |
| Emotional Mental Disorder | 0.713 | 0.048 | 216.699 | <0.001* | 2.041 (1.856-2.244) |
| Instant Food Consumption | 0.102 | 0.040 | 6.722 | 0.010* | 1.108 (1.025-1.197) |
| Constant | -3.844 | 0.113 | 1165.952 | <0.001 | 0.021 |

\* = *p-value* <0.05

Through binary logistic regression analysis, this study found that male (PR=1.225; 95% CI: 1.119-1.341), unmarried subject (PR=0.711; 95% CI: 0.556-0.908), low education level (PR=1.286; 95% CI: 1.195-1.384), unemployed subject (PR=1.267; 95% CI: 1.166-1.377), BMI above average (PR=2.472; 95% CI: 2.295-2.662), diabetes mellitus subject (PR=3.688; 95% CI: 2.630-5.173), emotional mental disorder subject (PR=2.041; 95% CI: 1.856-2.244), frequent consume instant food (PR=1.108; 95% CI: 1.025-1.197), all together were linked to hypertension among young adults.

## Discussion

Most young adults are unaware of their abnormally high blood pressure and only realize its effects when they age (Gooding et al., 2014). Young adults are less sensitive to hypertension than older people because young adults visit doctors less often than older people (Everett & Zajacova, 2015). There is a long-term impact of early-life risk factors of hypertension in young adults. Hypertension patients aged below 40 years have low awareness, low diagnosis, and poor blood pressure control (Johnson et al., 2014). This study provided information regarding the risk factors related to hypertension.

Among 73,800 subjects, the prevalence of hypertension among young adults in this study is 4.1% based on RISKESDAS 2018 data. The prevalence of hypertension among young adults in this population is lower than in a study in Zimbabwe, i.e., 7.4%, based on survey conducted in Harare, Bulawayo, and Mashonaland East (Sabapathy et al., 2023). Similarly, a study conducted in Lahore, Pakistan showed that the prevalence of hypertension among young adults was 15.7% (Zaib Zahid et al., 2023). The study conducted in India has a higher prevalence of hypertension among young adults, i.e., 13.8%, based on The National Family Health Survey-4 data (Krishnamoorthy et al., 2023).

Males are more likely to suffer from hypertension than females (Ahammed et al., 2021; Liu et al., 2017; Singh et al., 2017; Wyszyńska et al., 2023). Low education levels are more likely to suffer from high blood pressure (Mohammed Nawi et al., 2021; Singh et al., 2017; Wyszyńska et al., 2023). Marital status affects the occurrence of hypertension, i.e., an unmarried subject has a greater risk of hypertension (Ramezankhani et al., 2019; Singh et al., 2017; Thawornchaisit et al., 2013). Unemployed subjects risk developing hypertensive disease more than employed people (Kato et al., 2023; Mohammed Nawi et al., 2021; Rismadi et al., 2021). Having exceeded body mass index is more at risk of developing hypertension (Ondimu et al., 2019; Patil et al., 2017; Thawornchaisit et al., 2013; Wyszyńska et al., 2023; Zaki et al., 2021). People with diabetes mellitus are more at risk of developing hypertension (Singh et al., 2017; Thawornchaisit et al., 2013; Tsimihodimos et al., 2018; Zaki et al., 2021). Emotional mental disorder is one of the conditions that promotes the development of hypertension (Alwani et al., 2023; Hu et al., 2015; Munakata, 2018; Ojike et al., 2016). Smoking habits have a significant influence on hypertension (Jang, 2021; Lusno et al., 2020; Maharjan, 2018; Meher et al., 2023; Saladini et al., 2016; Wang et al., 2020). Frequently eating instant sodium-rich foods also affects hypertension (Alsabieh et al., 2019; De Deus Mendonça et al., 2017; Shi et al., 2019).

Males are more at risk than females for hypertension. The results of this study are in line with the study in Albania regarding risk factors for hypertension among young adults, which states that males are more at risk of hypertension (Ahammed et al., 2021), and a study in Poland also showed similar results (Wyszyńska et al., 2023). A cross-sectional study conducted in Varanasi stated that males are at 1.97 times greater risk of hypertension (Singh et al., 2017). A study of young adults in Zimbabwe also found that males were 1.53 times more likely to develop hypertension than females (Sabapathy et al., 2023). Generally, males have less favorable habits compared to females, such as excess consumption of instant food, smoking, and alcohol consumption, leading to hypertension (Ahammed et al., 2021; Thawornchaisit et al., 2013). The low number of male visits to health services is also the reason for the higher rate of hypertension in males. Females are more likely to visit health services for consultation regarding birth control and regular gynecological services, where every visit will check their blood pressure so that females’ blood pressure is more controlled compared to males (Everett & Zajacova, 2015).

Unmarried subjects are more likely to suffer hypertension. The result was in line with a study conducted in Iran that showed unmarried subjects had a 1.55 higher risk of hypertension (Ramezankhani et al., 2019). The study conducted in Poland showed that married men had 1.58 more significant risk of hypertension than unmarried men (Lipowicz & Lopuszanska, 2005). Being married may be a protective factor against hypertension. Marriage is associated with a greater likelihood of lower nighttime systolic blood pressure (SBP) in men. Marriage might had some advantages, i.e., better health, good nutritional, and maintained psychiatric wellness (Causland et al., 2014). The treatment for hypertension among young adults was better in married men. Married improved men’s compliance against medical treatment (Kim & Kim, 2020).

Education affects the incidence of hypertension. According to a study conducted in China, education limited to elementary school or below has a greater risk of hypertension compared to someone who graduated from middle school or above (Sun et al., 2022). A low education level has an impact on low knowledge, and this affects a person’s behaviour. Education influences a person’s behaviour in receiving and implementing information. There is a relationship between education and the occurrence of hypertension. Highly educated individuals have easier access to health information. A person’s education level influences the quality of behaviour in receiving and processing information, which will impact their health status (Pebrisiana et al., 2022; Singh et al., 2017).

This study found working status has a significant influence on hypertension, whereas unemployed have a greater risk of hypertension. This study’s results align with a systematic review and meta-analysis study in Southeast Asian Countries, which states that the unemployed have a greater risk of hypertension (Mohammed Nawi et al., 2021). Unemployed people are 1.39 more at risk of hypertension compared to employed people (Blok et al., 2022). The results of a nationwide study in China state that unemployment conditions are a high risk of hypertension because not working will affect economic capacity. Thus unemployment will indirectly make someone less aware of hypertension treatment and control (Zheng et al., 2020). Unemployment is more at risk of hypertension because of low income; a person will have a limited variety of food, and health services received are more limited (Rismadi et al., 2021).

This study shows that body mass index has a significant effect on hypertension. Having a BMI≥25 has a 2.47 times greater risk of hypertension compared to having a BMI<25. Some studies show similar results, including a study in Poland which states that having a BMI above average is a greater risk of hypertension (Wyszyńska et al., 2023), a study conducted in Nagpur (Patil et al., 2017), and the study conducted on South Asian population-based from nationally representative surveys (Shahimi et al., 2022). A cohort study conducted in Thailand over four years showed that BMI was significantly associated with an increased risk of hypertension (Thawornchaisit et al., 2013). A study conducted among young adults in Tenwek Mission Hospital, Bomet County, Kenya, in 2018 stated that having a BMI≥25 had a 3.05 more significant risk of hypertension (Ondimu et al., 2019). A study conducted in Malaysia stated that obese people have a 4.29 times risk of developing hypertension (Zaki et al., 2021). Likened to people with normal BMI, people with BMI≥25 are more likely to have metabolic disorders, such as fat metabolism disorder and insulin antibody disorder. Metabolic disorders can trigger cardiovascular disease, one of which is hypertension (Tang et al., 2022). Apart from other risk factors, BMI directly affects blood pressure. Nevertheless, the mechanism of BMI’s influence on blood pressure is not definite (Landi et al., 2018). Several studies show that BMI is a critical and sensitive indicator for predicting high blood pressure (H. A. Lee & Park, 2018; Wu et al., 2019)

Comorbidities, such as diabetes mellitus, result in increased blood pressure. This study showed that diabetes mellitus has a significant impact on hypertension. A study conducted by (Petrie et al., 2018) indicates that the presence of comorbid diabetes mellitus puts a person at twice the risk of hypertension. A prospective cohort study conducted over four years in a national Thai Cohort Study showed similar results, where diabetes mellitus was strongly associated with hypertension (Thawornchaisit et al., 2013). Diabetes mellitus is also a variable that affects hypertension, as shown by a study conducted in Malaysia (Zaki et al., 2021). Research conducted in South Korea through The Fourth Korea National Health and Nutrition Examination Survey (KNHANES IV) states that hypertension is more prevalent in adults with diabetes mellitus in the general population and for both genders (H. S. Lee et al., 2013). Hypertension occurs due to complex metabolic disorders caused by diabetes mellitus. Increased circulating fluid volume and peripheral vascular resistance in patients with diabetes mellitus occur due to insulin resistance, which increases body fluid volume. This condition is closely related to increased blood pressure (Ohishi, 2018).

Psychological conditions also influence cardiovascular diseases, including hypertension. Psychologically, distress that occurs causes an increased risk of hypertension, 1.53 greater than a person without a mental disorder (Ojike et al., 2016). Stress was significantly related to hypertension in a cross-sectional study in China, where stressful conditions put a person 1.2 times more at risk of hypertension (Hu et al., 2015), and the results of a study in Jakarta, Indonesia showed that mental-emotional disorders made the risk of hypertension 4.095 times greater (Ekaningrum, 2021). Emotional mental disorders, e.g., stress, will trigger endothelial dysfunction that allows for an increase in blood pressure (Munakata, 2018). Emotional mental disorders may trigger habits that can cause an increase in blood pressure, such as physical inactivity due to a lack of enthusiasm for daily activities, smoking, consuming alcohol, and eating instant foods that contain high amounts of sodium.

This study found that instant food consumption leads to hypertension condition. A prospective cohort study in Spanish stated that high consumption of ultra-processed food creates a greater risk of hypertension (De Deus Mendonça et al., 2017). An 8-year cohort study in Thailand showed that dietary patterns are associated with hypertension. An unhealthy diet includes consuming processed foods, including instant foods that are low fiber, high in carbohydrates, saturated fat, and high in sodium (Shi et al., 2019). Generally, the combinations contained in instant food are carbohydrates, fat, and salt (Thawornchaisit et al., 2013). Frequent consumption of instant food will trigger cardiometabolic, which causes an increase in blood pressure (Shin et al., 2014). People often consume high-sodium instant foods. If high amounts of sodium are consumed continuously, it will lead to hypertension (Ekaningrum, 2021).

## Conclusion

The prevalence of hypertension among young adults in this study was 4.1% based on RISKESDAS 2018 data. Gender, education levels, working status, body mass index, diabetes mellitus, emotional mental disorder, and instant food consumption were associated with hypertension among young adults in Indonesia. The results of this study can used to predict risk factors for hypertension among young adults in Indonesia. Preventive efforts against hypertension in Indonesia should be performed by educating the public regarding behaviors that trigger hypertension so that modifiable risk factors will be more controlled. Early detection of health among young adults also needs to be improved, considering the low awareness and willingness to go to health services. This study has limitations, so the recommendation for future researchers is to examine more variables that may become a risk for hypertension.

## Data Availability

All data produced in the present study are available upon reasonable request to the authors

